# Which Office-Based Cardiovascular Risk Score is Suitable for Pokhareli Nepalese: Globorisk, WHO CVD, or Framingham?

**DOI:** 10.1101/2025.09.17.25336000

**Authors:** Grishma Adhikari, Shweta Ranabhat, Bipin Sapkota, Chiranjivi Adhikari

## Abstract

**Introduction:** Cardiovascular diseases (CVDs) are the leading cause of death. In resource-limited settings, office-based methods offer cost-effective alternatives to laboratory-based methods for primary prevention. Although the WHO Charts are used in the PEN (Package of Essential Non-communicable Diseases) program, their predictability is limited. In Pokhara Metropolitan, Nepal, this study compared the risk scores and agreement levels of three office-based CVD risk prediction models—WHO CVD, Globorisk, and Framingham Risk Score (FRS).

**Methods:** A community-based, cross-sectional study was conducted among 532 individuals (≥30 years) from Pokhara. Sociodemographic data and risk factors were collected through interviews, and anthropometric measurements. Descriptive statistics, t-tests, ANOVA, and Cohen’s kappa were used to compare risk categories and agreement levels, while linear regression analyzed trends across algorithms.

**Results:** CVD risk estimates differed significantly: WHO CVD (4.51 ± 3.46), Globorisk (7.35 ± 7.14), and FRS (9.59 ± 8.34), (F-statistics = 78.04, p <.01). Globorisk and the FRS showed very good model fits (R²= 85% and 90% respectively), whereas the WHO CVD showed an excellent model-fit (R²= 94%), doubting overfitting. The level of agreement was fair between Globorisk and WHO CVD (Kappa = 0.327, p <.01), slight between WHO and FRS (Kappa = 0.19, p < .01), and moderate between Globorisk and FRS (Kappa = 0.48, p < .01). The highest agreement was between Globorisk and FRS, particularly for females, and the lowest between WHO and FRS, especially for males. Ethnicity, education, marital status, and socio-economic factors were associated with CVD risk.

**Conclusion:** CVD risk predictions varied, with FRS predicting the highest risk and WHO CVD the lowest; with a difference of more than five percent. Variation was highest in low-risk (23.9%) and lowest in moderate-risk (10.4%). Ethnicity, occupation, education, marital status, and socio-economic factors should be considered before using appropriate algorithms.

**SUMMARY:** *What is already known on this topic:* WHO/ISH 2007 (World Health Organization/International Society of Hypertension) cardiovascular disease (CVD) risk charts are currently used in Nepal’s Primary Health Care Centers. These charts have been updated in 2019 (as WHO CVD 2019 risk charts). However, these both charts still have limitations, particularly in underestimating the risk.

*What this study adds:* This study compared risk scores provided by WHO CVD with other non-laboratory algorithms like Globorisk and the Framingham Risk Score (FRS). Our analysis included Cohen’s kappa statistics to find the agreement level among the risk categories and found differences in the mean risk scores among them. Ethnicity, educational level, and the marital status should also be considered when assessing CVD risk.

*How this study might affect research, practice, or policy:* Our findings highlight the variability in CVD risk predictions, with Globorisk demonstrating the most consistent risk estimates, while FRS overestimated and WHO CVD underestimated moderate and high risk. Globorisk may be a better predicting office-based model for Pokhareli Nepalese.

## INTRODUCTION

Cardiovascular diseases (CVDs), primarily heart attacks and strokes, account for one-third of all global deaths.^[1]^ CVD deaths doubled from 1990 to 2019 and may reach 23 million by 2030.^[2,3]^ Over three-quarters of these fatalities occur in Low- and Middle-Income Countries (LMICs). ^[1]^

The Sustainable Development Goal (SDG) 3.4 targets a one-third reduction in premature non-communicable diseases (NCDs) deaths by 2030.^[3,4]^ In Nepal, NCDs cause 66% of deaths, with CVD accounting for 44% of NCD-related fatalities, mostly affecting individuals aged 30-69. ^[3,5]^ CVD risk assessment relied on lab-based algorithms, while non-laboratory algorithms yield comparable outcomes, are cost-effective, and protect low-risk individuals from unnecessary harm.^[6,7]^

Several studies show discrepancies in 10-year CVD risk classifications. In Bangladesh, one-third of individuals were classified as low risk by WHO/ISH (World Health Organization/International Society of Hypertension) risk chart being categorized as moderate risk by Globorisk and FRS (Framingham Risk Score). Fair agreement was seen between WHO/ISH and Globorisk (kappa = 0.37), and between Globorisk and FRS (kappa = 0.34); however, agreement between WHO/ISH and FRS (kappa = 0.09)(p< .01)was minimum.^[8]^ Similarly, a study in Karachi, found moderate agreement (kappa=0.50) between Globorisk and FRS, with Globorisk classifying nearly half and FRS identifying one-third of newly diagnosed metabolic syndrome patients as moderate-to-high risk for 10-year CVD.^[9]^ In the Fasa Cohort Study in Iran, the mean scores for the non-laboratory-based WHO CVD and Globorisk risk models were 7.2 ± 4.9 and 6.2 ± 6.4, respectively, with substantial agreement between the two (kappa = 0.78).^[10]^ Furthermore, study in Srilanka, showed that the agreements between FRS BMI (Body Mass Index)-based and WHO/ISH without-cholesterol models were fair (kappa= 0.234, p< .01). FRS risk predictions were higher than those of the WHO/ISH charts, concluding that these two models should not be used interchangeably when studying South Asian populations.^[11]^

In Nepal, WHO/ISH charts are used in the health system applied by PEN (Package of Essential Non-communicable Diseases) Package. ^[12]^ Although the WHO Chart is used in the Nepalese Health System, its consistency with other non-laboratory risk charts hasn’t been evaluated. CVD risk is calculated and compared using three algorithms: WHO CVD^[13]^, Globorisk ^[10]^ and Framingham risk score (FRS)^[14,15]^ in residents of Pokhara Metropolitan. In this context, we aimed to evaluate the consistency and agreement of these three risk charts, we hope to get insights into the result consistency of the model.

WHO/ISH non-laboratory algorithms were developed using hypothetical datasets for six WHO regions based on risk factor prevalence. The internal validation of the WHO/ISH non-laboratory based algorithms was not reported.^[16]^ A cross-sectional study in Nepal in 2023, using WHO/ ISH risk chart suggests potential overestimation by it, suggesting the need for further validation in resource-limited settings.^[17]^

Similarly, WHO CVD 2019 charts were derived from 85 cohorts, mostly from high-income countries. To recalibrate the models for LMIC data were used from the Global Burden of Disease project; however, the data do not have country-specific disease risk estimates. It categories fewer individuals as high risk.^[18]^ This tool underestimates CVD risk among those with target organ damage, and diabetic patients with complications.^[19]^

This study aimed to calculate and compare CVD risk using three algorithms: WHO-CVD, Globorisk, and FRS. Additionally, we examined the socio-economic factors influencing CVD risk.

## METHODS AND MATERIALS

### Study Design, Population, and Setting

We conducted a community-based cross-sectional analytical study among adults aged 30 years and above in Pokhara Metropolitan. According to the 2021 census, Pokhara spans 464.24 sq km with 513,504 people, a density of 1,106 per sq km, 140,459 households, a sex ratio of 93.04 males per 100 females, and an 88.7% literacy rate (94.2% males, 83.7% females).^[20,21]^

### Sample size and Sampling technique

We calculated sample size using Cochran’s formula assuming 13.6% prevalence of moderate to high risk of CVD in 10 years, 95% confidence interval, 5% absolute error and 2.5% design effect. The calculated sample size was 532.^[22]^

### Data collection

Data was collected by trained interviewers through face-to-face interviews using the Solstice mobile app (August–September 2023) with a structured questionnaire covering socio-demographics, anthropometry, blood pressure, and risk prediction tools.

### Socio-demographic characteristics

It includes age (in years), sex (Male/Female), ethnicity (Brahmin or Chhetri/Janajati/Dalit/ Madhesi/ Other), Marital status (Married/ Never Married/ Widowed/ Separated/ Divorced), SES (Socio-economic Status) (Upper/Upper Middle/Lower Middle/Upper Lower/Lower). SES was assessed using Kuppuswamy scale.^[23,24]^ It had the components Employment status (Unemployed/Unskilled worker/Semi-skilled worker/Skilled worker/Arithmetic skill job/Semi-professional/Professional); Educational status (Illiterate/Literate, less than Middle school certificate/Middle school certificate/High school certificate/Higher secondary certificate/Graduate degree/ post-graduate or professional degree); Monthly family income characterized as (<4850/ 4851-14550/ 14551-24350/ 24351-36550/ 36551-48750/ 48751-97450/ >= 97451).^[24]^

### Anthropometric measurements

We measured height using calibrated stadiometer and weight using bathroom weighing scales. The least count of stadiometer and weighting scale were 0.1 cm and 0.1 kg respectively. We used standard protocol to measure weight and height.^[25]^

### Blood pressure (BP) measurement

BP was measured using an Omron sphygmomanometer following standard protocols to ensure accuracy, with a 2–3-minute interval between readings. ^[26]^

### Risk prediction tools

The main outcome of 10-yr CVD risk includes the risk of both fatal and nonfatal cardiovascular events, such as coronary heart disease (CHD), stroke (ischemic and hemorrhagic), myocardial infarction, coronary insufficiency, angina, transient ischemic attack, peripheral artery disease, and heart failure.^[13,14]^ We used three NCDs risk prediction tools: Globo risk, b) World Health Organization CVD risk, and c) Framingham CVD risk prediction score.

WHO has developed tables that predict the risk of CVDs based on factors like age, sex, systolic blood pressure, cholesterol, and smoking for laboratory purposes. There is a second set of tables that use BMI instead of cholesterol for non-laboratory assessment. In this study, the non-laboratory WHO CVD risk prediction chart for SEAR D to determine the likelihood of cardiovascular events over the next ten years for people 40-79 years is used. The chart is a cost-effective tool to stratify the entire population using risk categories like Low risk (<5%) in green color, Moderate risk (5-<10%) in yellow color, High risk (10-<20%) in orange color, Very high risk (20-<30%) in red color, extremely high risk (>30%) in deep red color.^[13]^

Globorisk predicts 10 years’ fatal and non-fatal CVD comprised deaths from IHD (Ischemic Heart Disease), sudden cardiac death or stroke and nonfatal myocardial infarction and stroke^[6]^. The non-laboratory risk score included age, sex, smoking, blood pressure, and BMI. It has been validated and calibrated using data from 182 countries and its external validity has been examined in different regions. No risk grouping has not been done here like other CVDs risk prediction models.^[27,28]^ Globorisk calculator was used to assess the risk.

Similarly, CVD is defined by the FRS as a composite of CHD (coronary death, myocardial infarction, coronary insufficiency, and angina), cerebrovascular events (including ischemic stroke, hemorrhagic stoke, and transient ischemic attack), peripheral artery disease (intermittent claudication), and heart failure. The non-laboratory-based model is based on age, sex, SBP (Systolic Blood Pressure) and treatment status, current smoking, diabetes and, BMI. ^[27]^ 10-year CVD risks were categorized as low (<10%), moderate (10–<20%), and high (>20%).^[14]^

### Data processing and statistical procedure

Data that was entered through Solstice mobile application was exported to the Statistical Package for Social Sciences (SPSS) Version 20 for analysis. SPSS was used for data entry and cleaning to ensure consistency.

Descriptive statistics were used for summarizing the independent and dependent variables. The continuous variables were presented as means and standard deviations (SD). Normality of continuous variables was explored visually (histogram). Whereas categorical variables were presented as frequencies and percentages.

The independent samples t-test was used to compare the means of different variables between risk categories within each CVD risk assessment tool (Globorisk score, WHO CVD Risk, FRS). Similarly, an ANOVA test was used to compare the means of three groups to determine if there are any statistically significant differences among them.

Cohen’s kappa was used to measure the levels of agreement among the tools as it is used to measure either interrater or intrarater reliability. The Kappa results were interpreted as follows: Kappa values <0 represent no agreement, 0.01–0.20 represent none to slight agreement, 0.21–0.40 represent fair agreement, 0.41–0.60 represent moderate agreement, 0.61–0.80 represent substantial agreement, and 0.81–1.00 represent nearly perfect agreement.^[29]^ Similarly, a Chi-Square test for trends analysis was conducted to assess the distribution of variables in CVD risk groups, with linear regression analysis used to explore trends in the risk prediction performance of different algorithms. Equiplot, a statistical tool, was used to create a visual representation of CVD risk distribution across different age groups and socio-economic strata.

### Ethical statements

Ethical approval with reference number 013-080/81 was taken for this study from the Institutional Review Committee of Pokhara University based on its protocol and format. Written approval was obtained from the Department of Health, Pokhara Metropolitan. Prior to data collection, both written and verbal informed consents were obtained from each participant.

## RESULTS

### Descriptive statistics

Nearly half (53.2%) of the participants were male. Nearly two-thirds (65.4%) were Brahmin/Chhetri, followed by one-fourth Janajati (26.9%), and then Dalit, Madhesi, and others. The mean age of the participants was 47.82 ± 11.67 years. Regarding occupation, nearly one third of them were semi-skilled workers (37.8%), one fifth unemployed (20.1%), and 23.9% had education below class VIII. The highest proportion of employees, nearly one-third (30.1%), had a monthly family income of Rs 14,551–24,350, followed by one-fifth (24.6%) earning between Rs 24,351–36,550. The majority of the participants (41.7%) fell into the Upper-Lower class category, followed by one-third (30.8%) in the Lower-Middle class. Among the participants, it was found that most of them (91.7%) were married, with the mean marital years of 23.91 ± 13.13 years (Supplementary Table 1).

### Socio-economic characteristics of mean differences of CVD risk scores

CVD risks differed significantly across the three algorithms (FRS, WHO, and Globorisk) based on ethnicity, occupation, education, SES, and marital status. Brahmins and Chhetris had higher mean CVD risk scores across the three algorithms compared to the Janajati and underprivileged groups (p’s ≤ .02). Less skilled and unemployed individuals showed higher CVD risk than those in professional roles (p < .01). In the same line, people who are less educated had higher risks across all measures (p’s<.01). Although income levels did not show significant differences in mean scores (p’s > .05), individuals with lower SES had higher risks in Globorisk and WHO scores (p’s < .01). Moreover, never married, divorced, widowed, and separated had greater mean CVD risks across all scores as compared to married counterparts (p’s≤.02). Notably, FRS scores did not show any significant differences in means of people with different occupations (p=.77), and the lower and upper SES (p=.18) (Supplementary Table 2).

### Comparative assessment of the risk scores-Globorisk, FRS, WHO CVD

The mean risk scores observed in the three risk algorithms showed a significant difference (F(df); 78.037; p, < .01) (Table 3).

**TABLE 1:**
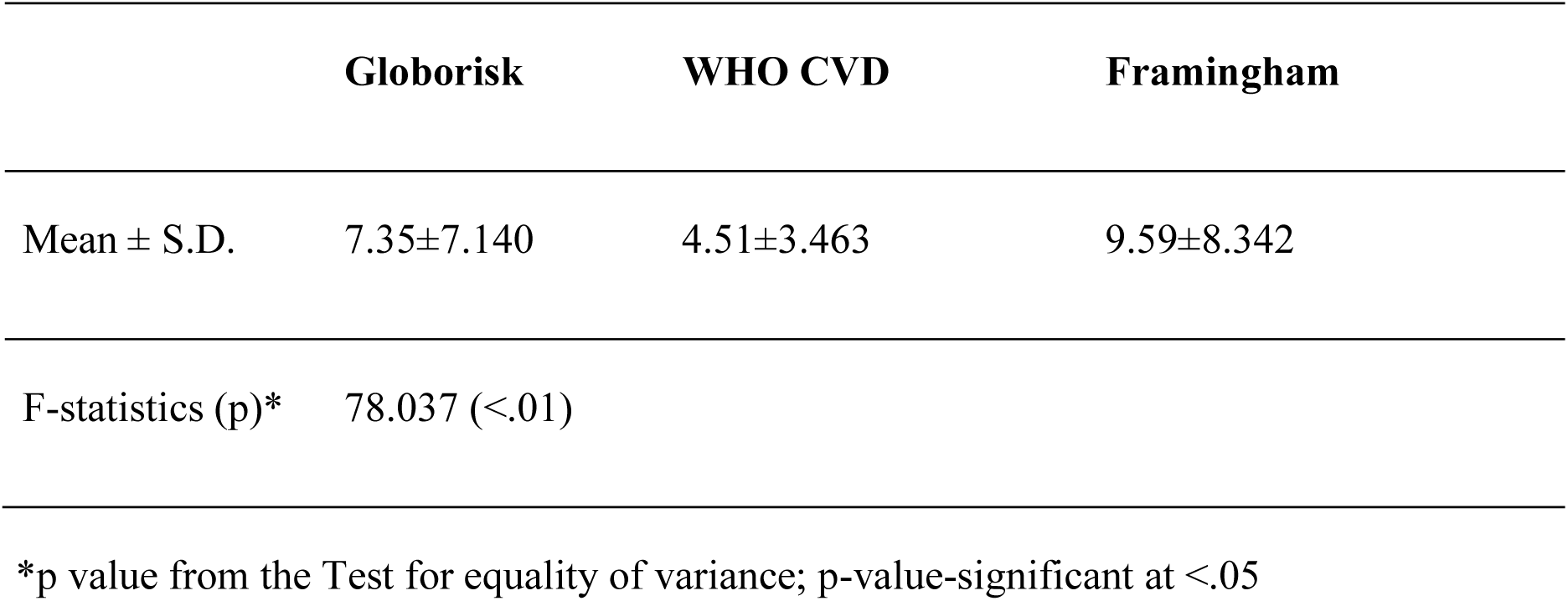
ANOVA TEST.

There was a fair agreement between Globorisk and WHO CVD (Kappa value of 0.33; p’s<.01). Similarly, there was a slight agreement between WHO and FRS (Kappa = 0.19; p’s<.01); which is the lowest. Globorisk and FRS showed the highest agreement, with a moderate agreement (Kappa = 0.48; p’s<.01). Globorisk and FRS showed the highest agreement, especially for females. WHO and FRS showed the lowest agreement, especially for males (Table 4).

**TABLE 2:**
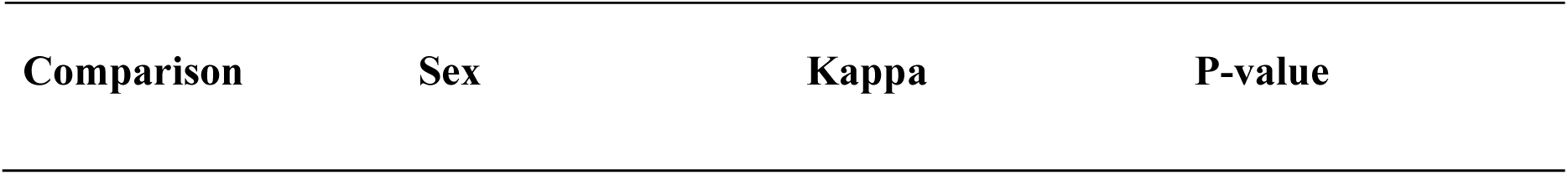
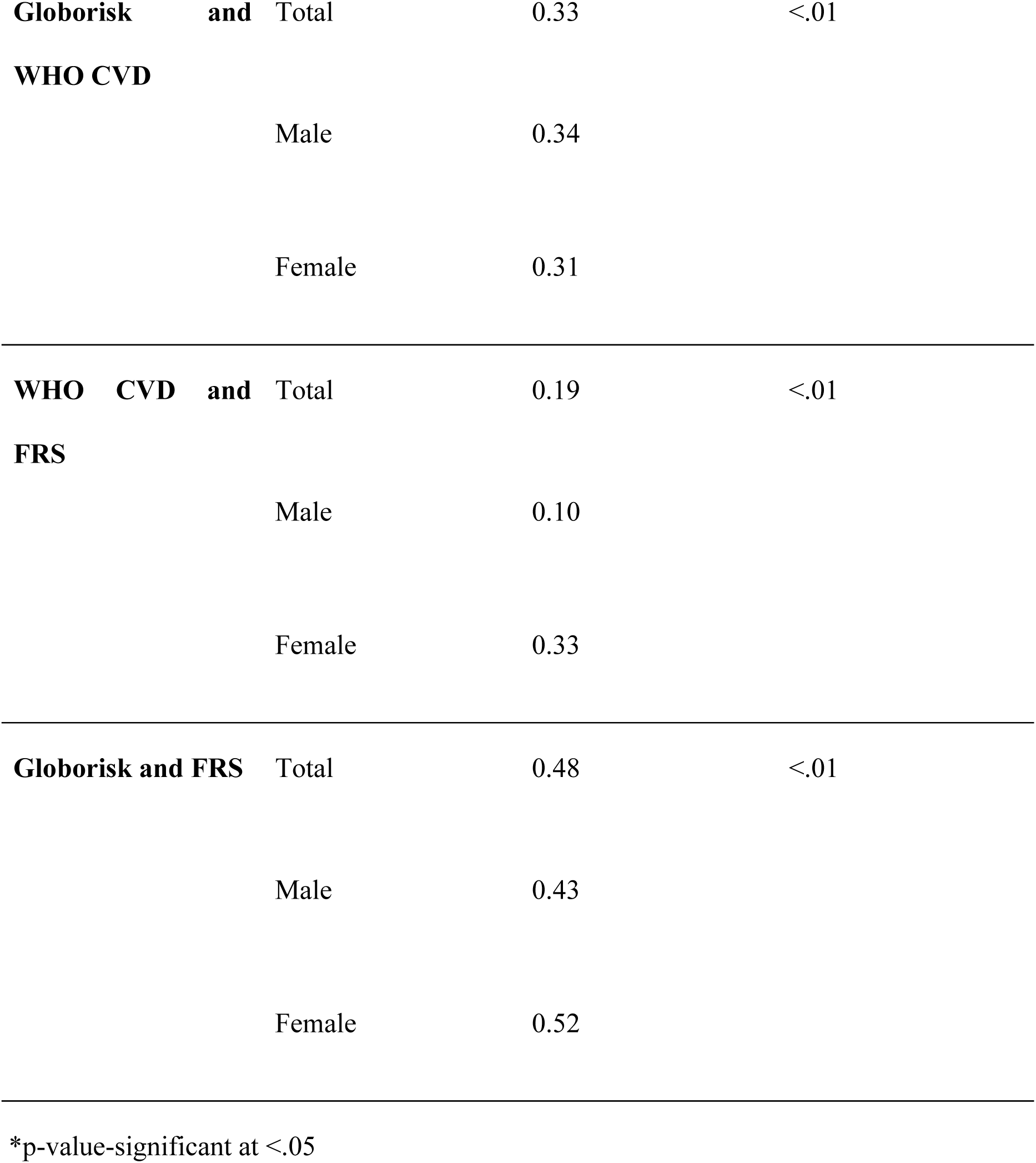
LEVEL OF AGREEMENT BETWEEN RISK SCORES.

The risk scores varied according to the value of CVD risk (p’s < .01). The trend was linear with a slope of 0.12 (p’s<.01), suggesting a significant linear relationship between the variables. The χ² test for non-linearity (p = .84) showed the absence of a significant non-linear relationship. The linear regression model demonstrated a strong fit, with a coefficient of determination (R²) of 80% or higher for all three risk scores. (Figures 1a and 1b)

**Figure 1:**
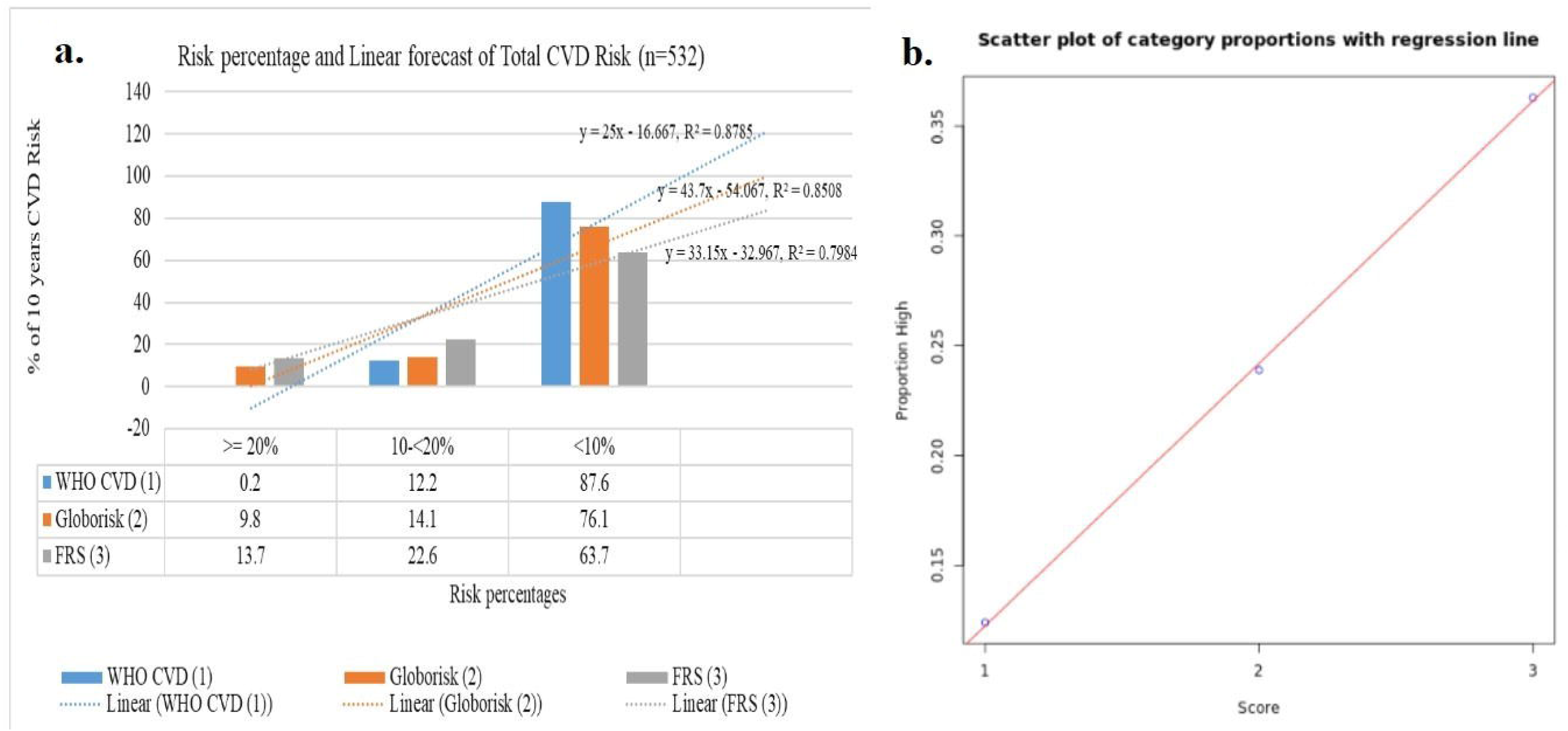
Comparative Linearities of Risk Categories of Three Algorithms. **Fig 1a.** χ² for trend (df), p, slope; 82.67 (1), <.01, 0.12. χ² for linearity (df), p; 0.04 (1), .84 **Fig 1b.** Scatter plot of high risk proportion across three risk scores with regression line (Calculated from counts, moderate and high risk counts merged).

### Comparative CVD Risk between Male and Female

The trend was linear with a slope of 0.16, (p <.01), indicating a significant relationship between the variables. The χ² test for non-linearity (p = 1) showed the absence of a significant non-linear relationship. The linear regression model demonstrated a strong fit, with a high coefficient of determination (R² > 85%) across all three risk scores: WHO CVD (R² = 0.94), Globorisk (R² = 0.85), and FRS (R² = 0.90) (Figure 2a and 2b).

**Figure 2:**
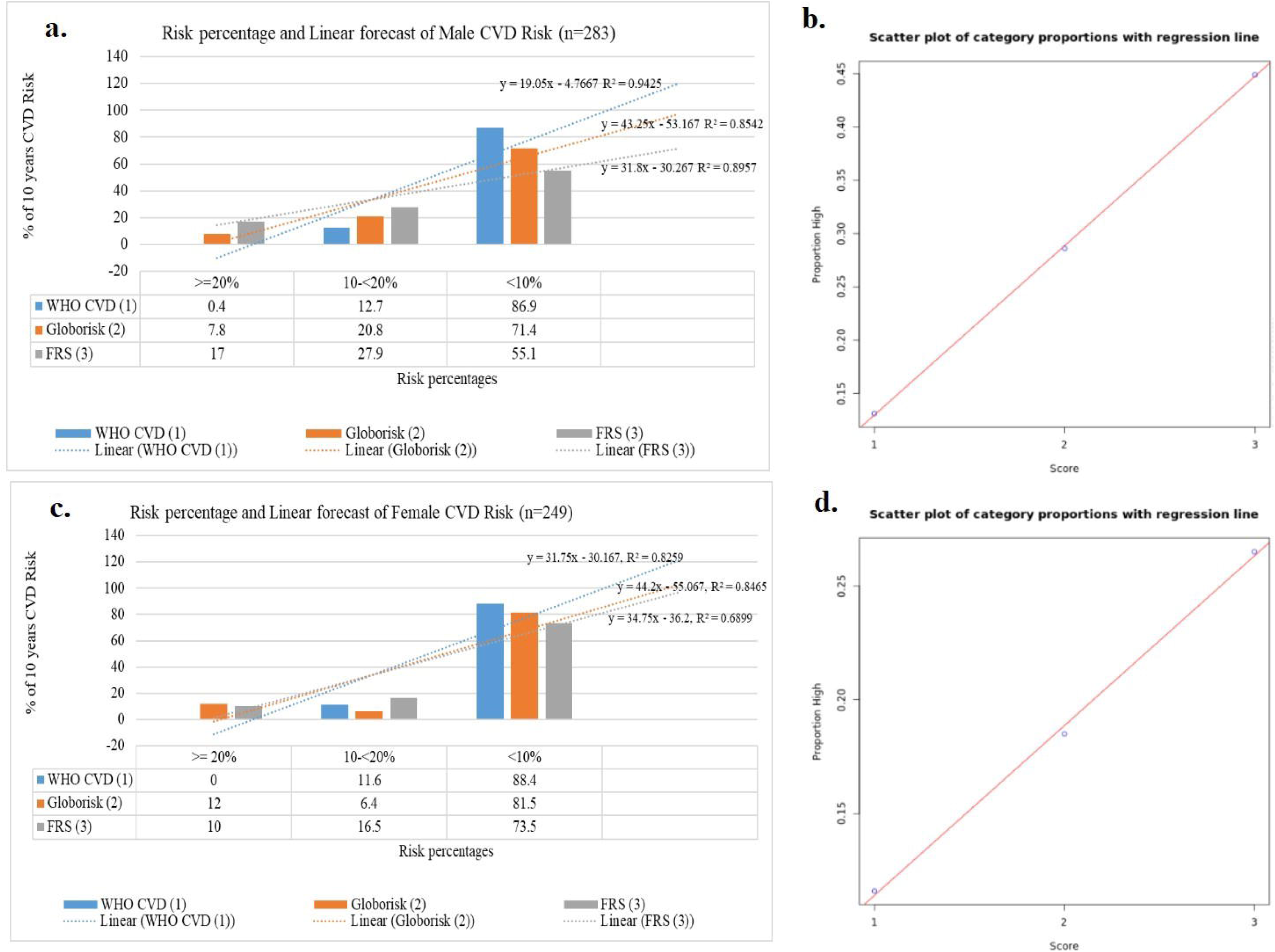
Comparative Linearities of Sex-Wise Risk Categories Across Three Risk Algorithms. **Fig 2a.** χ² for trend (df), p, slope; 69.71 (1), <.01, 0.16. χ² for linearity (df), p; 0.01 (1), .91. **Fig 2b.** Scatter plot of high risk proportion across three risk scores with regression line. **Fig 2c.** χ² for trend (df), p, slope; 17.95 (1), <.01, 0.07. χ² for linearity (df), p; 0.04 (1), .84. **Fig 2d.** Scatter plot of high-risk proportion across three risk scores with regression line. (Calculated from counts; moderate and high risk counts merged.)

The trend was linear with a slope of 0.07, (p’s= <.01), suggesting a significant linear relationship between the variables. The χ² test for non-linearity (p = .84) showed no significant non-linear relationship. The linear regression model demonstrated a strong fit, with a high Coefficient of Determination (R²) of 82.59% and 84.65%, for WHO CVD and Globorisk, respectively. However, the FRS model had a lower R² of 70%, suggesting a moderate fit for the linear regression model. (Figure 2c and 2d).

### Comparative CVD Risk across age and socio-economic status

WHO CVD risk distributed by age and stratified by sex and SES (n=532), showed the age-related variation in CVD risk scores using the WHO CVD algorithm. Risk scores were lower for younger individuals (∼30 years old) and increased with age, particularly in lower SES groups. This trend was especially visible in lower SES categories, where data points spread further to the right (higher risk scores) and vertically (older ages). (Figure 3a).

**Figure 3:**
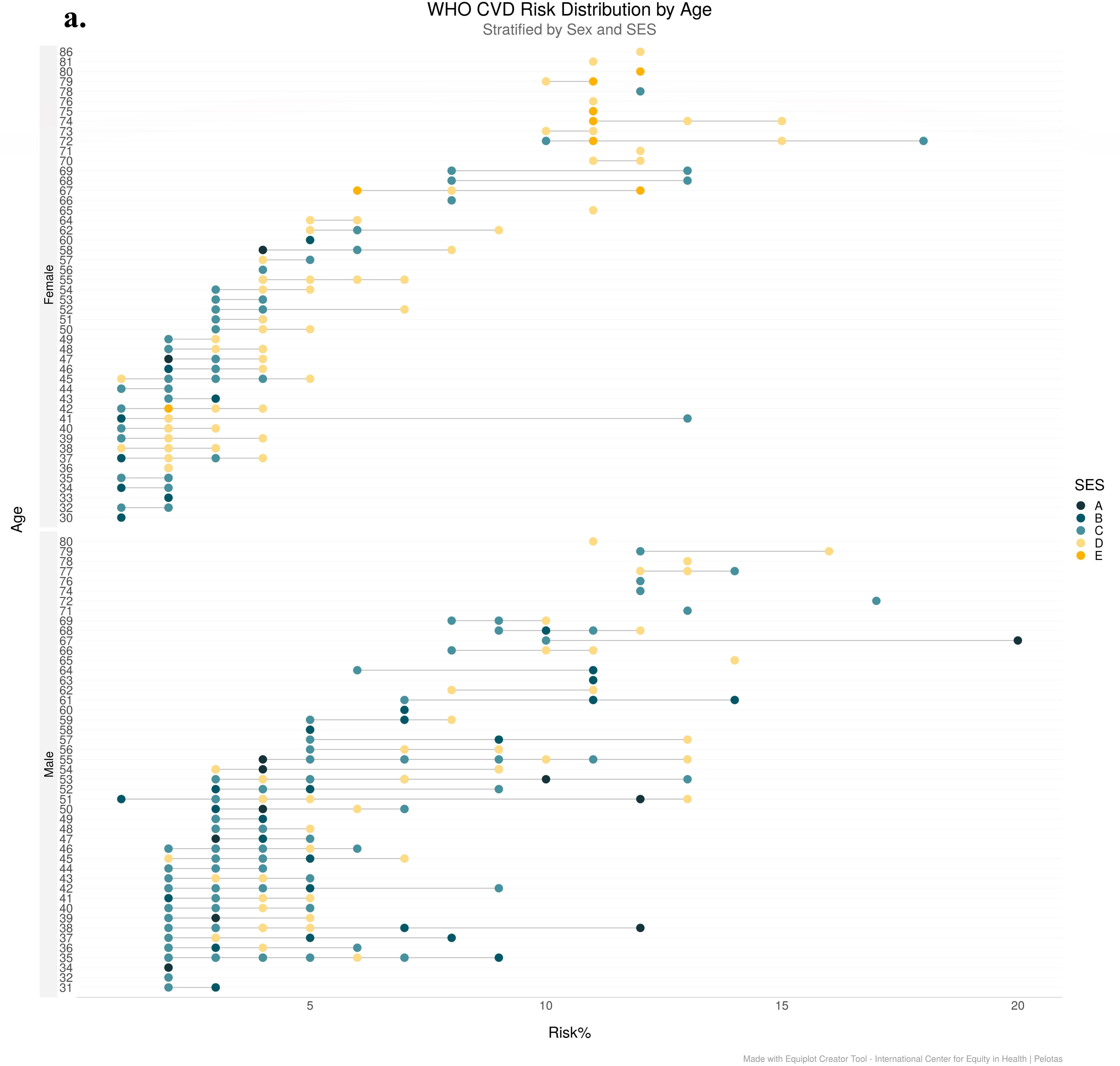
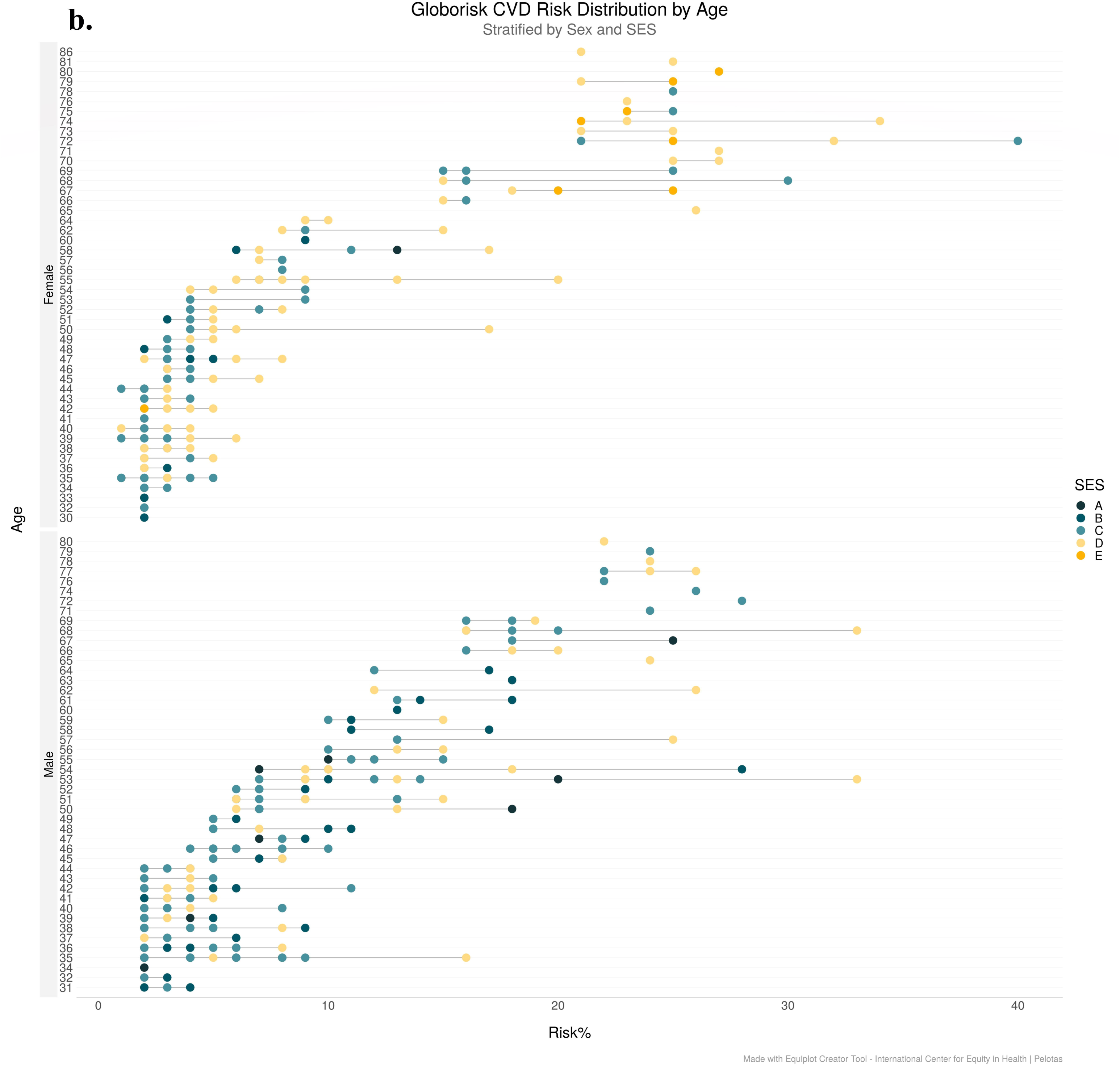
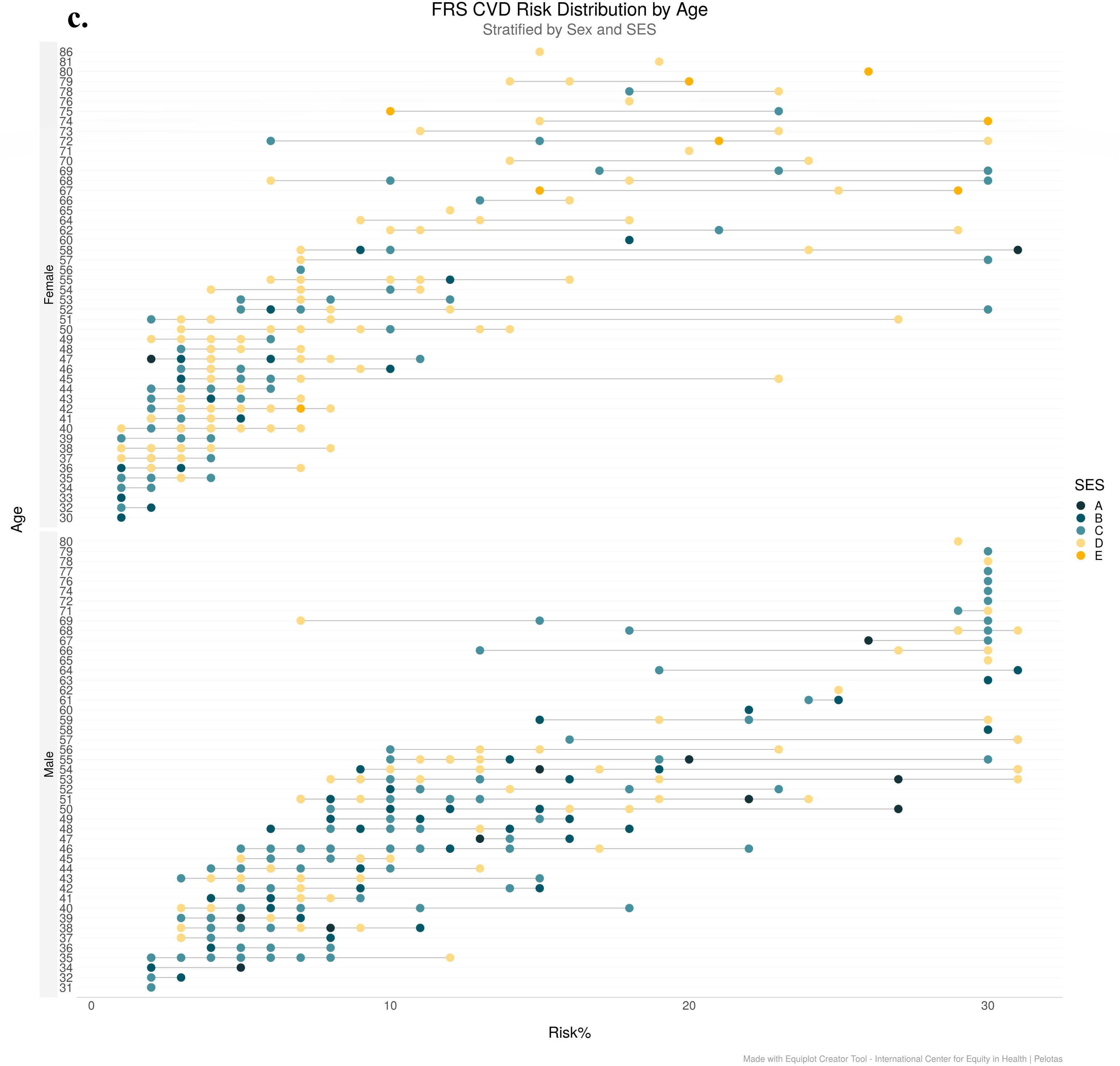
CVD Risk Scores Stratified with Age And Socio-Economic Status. **Fig 3a.** WHO CVD Risk Scores by Age with SES (n=532);SES, A-Upper, B-Upper Middle, C-Lower Middle, D-Upper Lower, E-Lower **Fig 3b.** Globorisk Scores by Age with SES (n=532);SESA-Upper, B-Upper Middle, C-Lower Middle, D-Upper Lower, E-Lower **Fig 3c.** FRS by Age with SES (n=532); SES,A-Upper, B-Upper Middle, C-Lower Middle, D-Upper Lower, E-Lower

Globorisk, distributed by age and stratified by sex and SES (n=532), showed that risk scores increase with age, particularly for lower SES groups. Older individuals and those from lower SES backgrounds exhibited higher risk scores (Figure 3b).

Consistently, FRS distributed by age and stratified by sex and SES (n=532), showed that people at older ages exhibited an upward trend in risk scores (Figure 3c).

## DISCUSSION

Office-based risk scores —WHO CVD 2019, Globorisk, and FRS revealed different proportions of moderate-high risks among different population segments and across socio-demographic characteristics. We discuss, first, the risk variation and its magnitude, across the globally validated three algorithms—WHO CVD, Globorisk, and FRS— the variation occurred among the male, female, and total Pokhareli population. We further explore the impact of socioeconomic factors, including ethnicity, occupation, and education, on CVD risks.

### Risk categories and its variation magnitude in three algorithms

High and moderate risk scores were observed differently across the three risk scores. The slope of total CVD risk across three risk scores is non-zero (0.12); suggesting that the three risk scores show inconsistencies, with a variation of 8% (R²’s; 0.80-0.88) across three risk scores. Consistently, a Burkina Faso study of 3081 participants found mean 10-yr risks of 2.5% by WHO, 4.6% by FRS, and 4.0% by Globorisk.^[27]^ Similarly, both Globorisk and FRS identified a greater higher proportion of individuals at moderate risk (37.0% and 38.8%, respectively) compared to the WHO/ISH risk charts, which classified only 15.3% of participants in this category. Similar trends were observed in the high-risk and very high-risk categories.^[8,18]^

Being in the same line of risk variation, we observed that the WHO CVD regressed with the greatest variation of effect (WHO CVD R²=0.88, Globorisk R²=0.85 and FRS R²=0.80) among the three, whereas the FRS gives a greater proportion of moderate and high risk (FRS=36.3%, Globorisk =23.9% and WHO CVD =12.4%). Similarly, a linear relationship (Δ >20%=13.5%, Δ 10-<20%=10.4%, <10%=23.9%) was observed with (χ²(df), p 0.04(1),.84), and a significant slope across three risk scores.

A CVD risk study in Bangladesh found higher CVD risk in males: 57.4% by FRS, 4.3% by WHO, and 33.3% by Globorisk. For females, FRS identified 16.4% at high/very high risk, compared to 4.2% by WHO and 4.8% by Globorisk. FRS identified a higher proportion of individuals at elevated risk, especially among males.^[8]^ This study too, for males, high and moderate risk scores were observed differently across 3 risk scores. The slope of CVD risk across 3 risk scores for males is non-zero (0.16); implying that the results given by 3 risk scores show inconsistent results, with a variation of 8.8% (R²’s; 0.85-0.94), across 3 risk scores. The greater amount of regression is shown by the WHO CVD risk score (WHO CVD R²=0.94, Globorisk R²=0.85 and FRS R²=0.90), while greater proportion of moderate and high risk is given by the FRS risk score (FRS=44.9%, Globorisk =28.6% and WHO CVD =13.1%). Similarly, a linear relationship was observed with (χ²(df),p 0.01(1),.91), and a significant slope across 3 risk scores. Likewise, for females, high and moderate risk scores were observed differently across 3 risk scores. The slope of CVD risk across 3 risk scores for males is non-zero (0.07); implying that the results given by 3 risk scores show inconsistent results, with a variation of 15.67% (R²’s;0.69-0.85), across 3 risk scores. The greater amount of regression is shown by the Globorisk score (WHO CVD R²=0.83, Globorisk R²=0.85 and FRS R²=0.70), while greater proportion of moderate and high risk is given by FRS risk score (FRS=26.5%, Globorisk =18.4% and WHO CVD =11.6%). Similarly linear relationship was observed with (χ²(df),p 0.04(1),.84), and significant slope across 3 risk scores. A 2015 Malaysian survey (n=8,253) also found higher CVD risk in males.^[31]^ With similar result, the study in Burkina Faso, the absolute 10-yr risk was seen higher among men than women across all three algorithms. It was seen to be 7.2% [CI 95%: 6.7-7.7] in men and 2.9% [CI 95%: 2.6-3.2] in women by FRS, 5.0% [CI 95%: 4.7-5.3] in men and 3.4% [CI 95%: 3.2-3.6] in women by Globorisk and 3.2% [CI 95%: 3.0-3.4] in men and 2.0% [CI 95%: 1.9-2.2] in women by WHO. Here, similar to our study

FRS gives the greatest proportion of high to moderate risk results, especially in men, with 7.2% classified as high risk, whereas Globorisk and WHO assign lower percentages to high or moderate risk.^[30]^

### Socio-economic factors influencing the risk variation

Brahmins and Chhetris have higher mean CVD Risk scores across the three algorithms-as compared to the Janajati and the Underprivileged groups (p’s≤.02). A cross-sectional study in Kathmandu, Nepal using WHO/ISH chart showed no significant difference in CVD risk across ethnic groups (p = .65). ^[32]^ But a study conducted in rural Nepalese population showed that Dalits are twice as likely (95% CI: 1.28–3.43) as Brahmin/Chhetri residents to have multiple CVD risk factors at the same time.^[33]^ People with lesser skilled jobs and the unemployed have higher mean CVD risks as compared to those with more professional roles, in Globorisk and WHO algorithms (p’s <.01). In contrast, a study in Kathmandu showed no association with occupational status (p = .25).^[33]^ Similarly, in contrast to our findings, regarding occupation, a higher risk of CVD was seen among paid workers as compared to unemployed individuals, with CVD risk of 5.4% [95% CI: 5.1-5.8] among paid workers and 3.8% [95% CI: 3.1-4.4] among jobless by FRS, 4.2% [95% CI: 4.0-4.4] among paid workers and 4.0% [95% CI: 3.5-4.5] among jobless by Globorisk and 2.7% [95% CI: 2.6-2.8] among paid workers and 2.4% [95% CI: 2.1-2.7] among jobless by WHO.^[30]^

People with lower education levels have higher risks across all measures (p’s < .01). This is supported by a study in Kathmandu, where education level significantly impacted CVD risk (p = .03); participants with no formal education had a (≥10%) greater risk (9.6%) than those with a Bachelor’s degree or higher.^[33]^ While in sharp contrast to our findings, a study in Burkina Faso showed that a higher degree of education was associated with high risk of CVD, with the risk of 4.9% [95% CI: 4.6-5.2] among unschooled and 6.1 % [95% CI: 4.4-7.8] among those with education higher than secondary by FRS, 4.1% [95% CI: 3.9-4.3] among unschooled and 4.8% [95% CI: 3.7-5.8] among those with education higher than secondary by Globorisk and 2.6% [95% CI: 2.5-2.7] among unschooled and 3.0% [95% CI: 2.3-3.6] among those with education higher than secondary by WHO.^[30]^ Although income levels do not show any differences in mean scores (p’s>.05) people with lower socio-economic statuses have higher risks with Globorisk and WHO scores (p’s<.01). A study in Bangladesh using the WHO/ISH, Globorisk, and FRS tools showed varying results based on income. WHO/ISH showed higher CVD risk for low-income individuals (4.4% at high/very high risk), while Globorisk and FRS found higher risk for high-income individuals (23.4% and 45.3%, respectively, at high/very high risk). Globorisk showed 12.6% and FRS showed 24.2% for low-income individuals at high/very high risk. ^[8]^ The findings of this study showed that never married, divorced, widowed, and separated individuals have higher mean CVD risks across all scores compared to their married counterparts (p’s ≤ .02). Consistently, a study carried out among elderly population in India using the WHO CVD risk charts showed that participants who were living alone, and widowed had 1.42, and 1.59 adjusted odds of having a high 10-yr CVD risk, respectively.^[34]^

In conclusion, the study found variability in CVD risk predictions. Globorisk demonstrated the most consistent risk estimates, while FRS overestimated and WHO CVD underestimated moderate and high risk. People belonging to the Brahmin/Chhetri ethnic group, unemployed, less educated, single, and individuals belonging to lower socio-economic class are at greater CVD risk (p < .05). (Figure 4)

**Figure 4:**
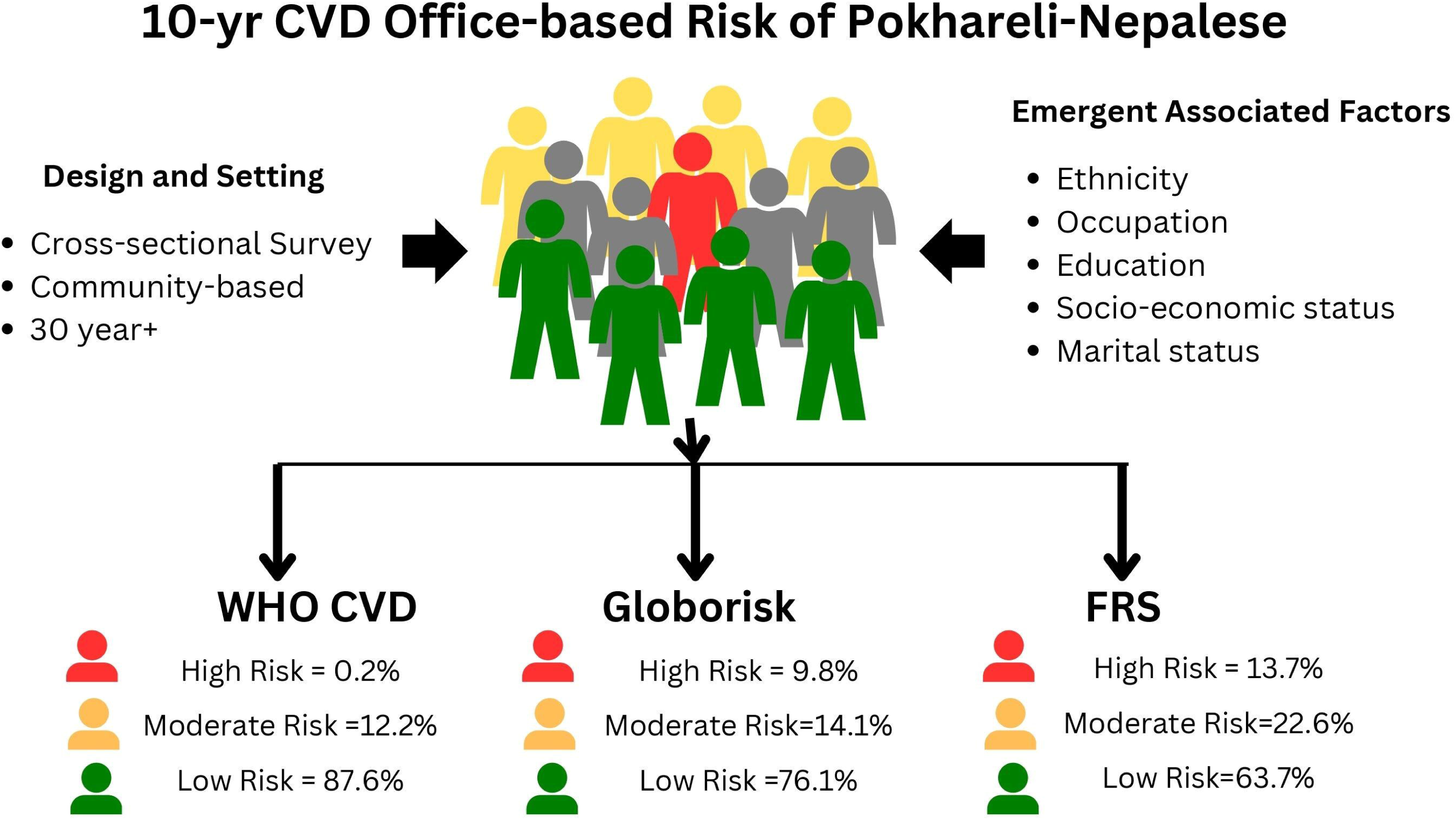
CVD risk estimates among Pokhareli-Nepalese (n=532) using three different risk prediction models: WHO CVD, Globorisk, FRS, along with associated risk factors (p < .05)

## STRENGTHS AND LIMITATIONS

The strengths of this study are that it is a community-based, cross-sectional analytical study providing a representative sample of the population in Pokhara Metropolitan, Nepal. The study compares three office-based CVD risk prediction models, the WHO CVD, Globorisk, and FRS, which are a significant strength. The analysis of socio-economic factors, such as ethnicity, education, occupation, and marital status, provides insights into the social determinants of health.

However, this study has some limitations like FRS risk score had to be converted into whole number, and values ≥30%, and values greater than 30% were not detailed specifically. Similarly, the weight of individuals ≤ 40 kgs was not accepted by the Globorisk calculator, so the minimum value had to be entered. Confidentiality was limited because data collection sites were open. The sample had over-representation of certain ethnic groups (e.g. Brahmin/Chhetri) and socioeconomic status (Upper Lower class) which may have skewed the findings. Furthermore, the study was conducted in a localized setting (Pokhara Metropolitan), which may limit the generalizability of findings to a broader population. The sample size of 532 is relatively small, which may limit the ability to validate the algorithms for Pokhreli population. Additionally, the WHO CVD showed an excellent model-fit (94%), doubting overfitting.

## Data Availability

Supplemental files of the journal.

## ABBREVIATIONS

BMI: Body Mass Index
BP: Blood Pressure
CHD: Coronary Heart Disease
CVDs: Cardiovascular diseases
FRS: Framingham risk score
ICD: International Classification of Diseases
IHD: Ischemic Heart Disease
ISH: International Society of Hypertension
LMIC: Low and Middle Income Countries
NCDs: Noncommunicable diseases
PEN: Package of Essential Non-communicable Diseases
SBP: Systolic Blood Pressure
SDG: Sustainable Development Goal
SEAR: South-East Asia Region
SES: Socio-economic Status
SPSS: Statistical Package for Social Sciences
WHO: WHO World Health Organization

## ACKNOWLEDGEMENT

We sincerely extend our gratitude to Assoc. Professor Dr. Niranjan Shrestha for reviewing the statistical part. We express our gratitude to students of Pokhara University who helped us in data collection—Bipin Adhikari, Sabnam Subedi, Mamata Basnet, Uday Singh, Suchita Subedi, Anita Tiwari, Isha Regmi, Shila GC, Sapana Subedi, Suraj Chaurasiya, Prachi Singh Gupta, Niruta Timalsina, Isha Regmi, Bina Gautam, Sabi Aryal, Sandesh Ranabhat, Sushant Lamichhane, Sujan Kandel, Anmol Bhattarai, Monika Gurung and Kundan Karki. We are thankful to all participants for their valuable time and participation in the study.

## REFERENCES

1. World Heart Federation. World Heart Report 2024 [Internet]. Available from: https://world-heart-federation.org/resource/world-heart-report-2024/ [cited 2024 Jun 2].

2. Roth GA, Mensah GA, Johnson CO, et al. Global Burden of Cardiovascular Diseases and Risk Factors, 1990–2019. J Am Coll Cardiol [Internet]. 2020 [cited 2024 Jun 2];76(25):2982–3021. Available from: https://www.ncbi.nlm.nih.gov/pmc/articles/PMC7755038/

3. WHO. NCD STEPS Survey 2019. Available from: https://www.who.int/docs/default-source/nepal-documents/ncds/ncd-steps-survey-2019-compressed.pdf [cited 2024 Jun 2].

4. World Health Organization. Implementation roadmap for accelerating the prevention and control of noncommunicable diseases in South-East Asia 2022–2030. 2022.

5. Ministry of Health and Population, Nepal. NBoD Report 2019 [Internet]. Available from: https://old.mohp.gov.np/attachments/article/449/5.%20NBoD%20Report%202019.pdf [cited 2024 Jun 2].

6. Jahangiry L, Dehghan A, Farjam M, et al. Laboratory-based and office-based Globorisk scores to predict 10-year risk of cardiovascular diseases among Iranians: results from the Fasa PERSIAN cohort. BMC Med Res Methodol. 2022;22(1):305.

7. Gaziano TA, Abrahams-Gessel S, Alam S, et al. Comparison of Nonblood-Based and Blood-Based Total CV Risk Scores in Global Populations. Glob Heart [Internet]. 2016 [cited 2023 May 30];11(1):37. Available from: https://globalheartjournal.com/article/10.1016/j.gheart.2015.12.003/

8. Mondal R, Ritu RB, Banik PC, et al. Cardiovascular risk assessment among type-2 diabetic subjects in selected areas of Bangladesh: concordance among without cholesterol-based WHO/ISH, Globorisk, and Framingham risk prediction tools. Heliyon. 2021;7(8):e07728.

9. Adil SO, Uddin F, Musa KI, et al. Risk Assessment for Cardiovascular Disease Using the Framingham Risk Score and Globorisk Score Among Newly Diagnosed Metabolic Syndrome Patients. Int J Gen Med [Internet]. 2023 [cited 2024 Nov 29];16:4295–305. Available from: https://www.ncbi.nlm.nih.gov/pmc/articles/PMC10518264/

10. Dehghan A, Rezaei F, Aune D, et al. A comparative assessment between Globorisk and WHO cardiovascular disease risk scores: a population-based study. Sci Rep. 2023;13(1):14229.

11. Mettananda KCD, Gunasekara N, Thampoe R, et al. Place of cardiovascular risk prediction models in South Asians; agreement between Framingham risk score and WHO/ISH risk charts. Int J Clin Pract [Internet]. 2021 [cited 2024 Nov 29];75(7):e14190. Available from: https://onlinelibrary.wiley.com/doi/abs/10.1111/ijcp.14190

12. World Health Organization. Package of essential non-communicable disease (PEN) intervention at primary health service setting: PEN training trainee’s manual. 2019 [cited 2024 Jun 12]. Available from: https://iris.who.int/handle/10665/310930

13. World Health Organization, Country Office for Nepal. Hearts: technical package for cardiovascular disease management in primary health care. 2020.

14. D’Agostino RB, Vasan RS, Pencina MJ, et al. General cardiovascular risk profile for use in primary care: the Framingham Heart Study. Circulation. 2008;117(6):743–53.

15. Framingham Heart Study. Cardiovascular Disease (10-year risk) [Internet]. Available from: https://www.framinghamheartstudy.org/fhs-risk-functions/cardiovascular-disease-10-year-risk/ [cited 2024 Jun 13].

16. Kariuki JK, Stuart-Shor EM, Leveille SG, et al. Evaluation of the performance of existing non-laboratory based cardiovascular risk assessment algorithms. BMC Cardiovasc Disord [Internet]. 2013 [cited 2023 May 29];13(1):123. Available from: 10.1186/1471-2261-13-123

17. Sitaula D, Dhakal A, Mandal SK, et al. Estimation of 10-year cardiovascular risk among adult population in western Nepal using nonlaboratory-based WHO/ISH chart, 2023: A cross-sectional study. Health Sci Rep [Internet]. 2023 [cited 2024 Dec 1];6(10):e1614. Available from: https://www.ncbi.nlm.nih.gov/pmc/articles/PMC10560824/

18. Islam JY, Zaman MM, Moniruzzaman M, et al. Estimation of total cardiovascular risk using the 2019 WHO CVD prediction charts and comparison of population-level costs based on alternative drug therapy guidelines: a population-based study of adults in Bangladesh. BMJ Open [Internet]. 2020 [cited 2024 Dec 1];10(7):e035842. Available from: https://www.ncbi.nlm.nih.gov/pmc/articles/PMC7371224/

19. Mendis S, Lindholm LH, Mancia G, et al. World Health Organization (WHO) and International Society of Hypertension (ISH) risk prediction charts: assessment of cardiovascular risk for prevention and control of cardiovascular disease in low and middle-income countries. J Hypertens. 2007;25(8):1578–82.

20. Central Bureau of Statistics. National Population and Housing Census 2021 Results [Internet]. Available from: https://censusnepal.cbs.gov.np/results/population?province=4&district=40&municipality=4 [cited 2024 Dec 15].

21. Pokhara Metropolitan City. पोखरा महानगरपािलका | गकी ⏢देश [Internet]. Available from: https://pokharamun.gov.np/ [cited 2024 Dec 15].

22. Khanal MK, Ahmed MSAM, Moniruzzaman M, et al. Total cardiovascular risk for next 10 years among rural population of Nepal using WHO/ISH risk prediction chart. BMC Res Notes. 2017;10(1):120.

23. Joshi SK, Acharya K. Modification of Kuppuswamy’s Socioeconomic Status Scale in the Context of Nepal, 2019. Kathmandu Univ Med J. 2019;17:1–2.

24. Sharma R. Revised Kuppuswamy’s Socioeconomic Status Scale: Explained and Updated. Indian Pediatr. 2017;54(10):867–70.

25. Luepker RV, Evans A, McKeigue P, et al. Cardiovascular survey methods. World Health Organization; 2004.

26. Omron Healthcare. User manual Omron BP742 (English - 28 pages) [Internet]. Available from: https://www.manua.ls/omron/bp742/manual?p=28 [cited 2024 Dec 17].

27. Hajifathalian K, Ueda P, Lu Y, et al. A novel risk score to predict cardiovascular disease risk in national populations (Globorisk): a pooled analysis of prospective cohorts and health examination surveys. Lancet Diabetes Endocrinol. 2015;3(5):339–55.

28. Ueda P, Woodward M, Lu Y, et al. Laboratory-based and office-based risk scores and charts to predict 10-year risk of cardiovascular disease in 182 countries: a pooled analysis of prospective cohorts and health surveys. Lancet Diabetes Endocrinol [Internet]. 2017 [cited 2024 Jun 25];5(3):196–213. Available from: https://www.ncbi.nlm.nih.gov/pmc/articles/PMC5354360/

29. McHugh ML. Interrater reliability: the kappa statistic. Biochem Medica. 2012;22(3):276– 82.

30. Cisse K, Samadoulougou S, Ouedraogo M, et al. Geographic and Sociodemographic Disparities in Cardiovascular Risk in Burkina Faso: Findings from a Nationwide Cross-Sectional Survey. Risk Manag Healthc Policy. 2021;14:2863–76.

31. Che Nawi CMNH, Omar MA, Keegan T, et al. The Ten-Year Risk Prediction for Cardiovascular Disease for Malaysian Adults Using the Laboratory-Based and Office-Based (Globorisk) Prediction Model. Med Kaunas Lith. 2022;58(5):656.

32. Dhungana RR, Khanal MK, Pandey AR, et al. Assessment of Short Term Cardiovascular Risk Among 40 Years and Above Population in a Selected Community of Kathmandu, Nepal. J Nepal Health Res Counc. 2015;13(29):66–72.

33. Khanal MK, Mansur Ahmed MSA, Moniruzzaman M, et al. Prevalence and clustering of cardiovascular disease risk factors in rural Nepalese population aged 40-80 years. BMC Public Health. 2018;18(1):677.

34. Loganathan V, Subramanian M, Sekhar Kar S. Ten-Year Risk for Developing Cardiovascular Disease Among Older Adults and Elderly in India: A Secondary Analysis of Wave-1 of Longitudinal Aging Study in India. Cureus [Internet]. 2024 Nov 30;15(10):e46772. Available from: https://www.ncbi.nlm.nih.gov/pmc/articles/PMC10632738/

