## Supplementary material for "Which Office-Based Cardiovascular Risk Score is Suitable for Pokhareli Nepalese: Globorisk, WHO CVD, or Framingham?": S1. Table 1. Table 2

### **SUPPLEMENTARY MATERIALS: TABLES**

**S1. Table 1: Socio-Economic Characteristics of The Participants**

| **Characteristics** | | **Frequency (n=532)** | | **Percentage** |
| --- | --- | --- | --- | --- |
| **Sex** | | | | |
| Male | | 283 | | 53.2 |
| Female | | 249 | | 46.8 |
| **Ethnicity** | | | | |
| Brahmin/Chhetri | | 348 | | 65.4 |
| Janajati | | 143 | | 26.9 |
| Dalit | | 29 | | 5.5 |
| Madhesi | | 7 | | 1.3 |
| Other (Dashnami and Thakuri) | | 5 | | 1.0 |
| **Occupation** | | | | |
| Unemployed | 107 | | 20.1 | |
| Unskilled worker | | 12 | | 2.3 |
| Semi-skilled worker | | 201 | | 37.8 |
| Skilled worker | | 46 | | 8.6 |
| Arithmetic skill job | | 80 | | 15.0 |
| Semi-professional | | 51 | | 9.6 |
| Professional | | 35 | | 6.6 |
| **Education** | | | | |
| Illiterate | | 64 | | 12.0 |
| Literate, less than Middle school certificate | | 127 | | 23.9 |
| Middle school certificate | | 86 | | 16.2 |
| High school certificate | | 124 | | 23.3 |
| Higher secondary certificate | | 72 | | 13.5 |
| Graduate degree | | 40 | | 7.5 |
| Post-graduate or professional degree | | 19 | | 3.6 |
| **Monthly Family Income** | | | | |
| <4850 | | 5 | | 0.9 |
| 4851- 14550 | | 21 | | 3.9 |
| 14551-24350 | | 160 | | 30.1 |
| 24351-36550 | | 131 | | 24.6 |
| 36551-48750 | | 73 | | 13.7 |
| 48751-97450 | | 94 | | 17.7 |
| >= 97451 | | 48 | | 9.0 |
| **Socioeconomic classes** | | | | |
| Upper | | 17 | | 3.2 |
| Upper Middle | | 121 | | 22.7 |
| Lower Middle | | 164 | | 30.8 |
| Upper Lower | | 222 | | 41.7 |
| Lower | | 8 | | 1.5 |
| **Marital Status** | | | | |
| Married | | 488 | | 91.7 |
| Never Married | | 9 | | 1.7 |
| Widowed | | 26 | | 4.9 |
| Separated | | 6 | | 1.1 |
| Divorced | | 3 | | 0.6 |
| **Other variables** | | ***(X̅ ± SD) [Md (Q1 ∼Q3)(Min∼Max)]** | | |
| Age | | (47.82 ± 11.67) years [45 (38∼53.75) (30∼ 86)] | | |
| Duration of conjugal years | | (23.91 ± 13.13)  years [22 (14.25∼30) (1∼68)] | | |

* X̅ , mean; SD, standard deviation; Md, median; Q1, first quartiles; Q3, third quartiles; Min, minimum value; Max, maximum value.

**S1.Table 2: Socio-Economic Characteristics of Mean Differences of CVD Risk Scores**

| **Variable** | **Globorisk score** | | | | **WHO CVD Risk** | | | | **Framingham Risk** | | | |
| --- | --- | --- | --- | --- | --- | --- | --- | --- | --- | --- | --- | --- |
|  | <10% | 10-20% | ≥ 20% | Mean±SD | <10% | 10-20% | ≥ 20% | Mean±SD | <10% | 10-20% | ≥ 20% | Mean±SD |
| **Ethnicity** | | | | | | | | | | | | |
| Brahmin/Chhetri | 247 (61.0) | 62 (82.7) | 39 (75.0) | 8.01± 7.46 | 296 (63.5) | 51 (78.5) | 1 (100) | 4.76± 3.73 | 200(59) | 92(76.1) | 56 (76.7) | 10.39± 8.45 |
| #Janajati | 158 (39.0) | 13 (17.3) | 13 (25.0) | 6.10±6.33 | 170 (36.5) | 14 (21.5) | 0 | 4.04±2.85 | 139(41) | 28 (23.3) | 17 (23.3) | 8.08±7.30 |
| X̅_d_ ± S.D. (S.E.); t (p) | 1.91±7.09 (0.65); 2.96 (<0.01) | | | | 0.72 ±3.45 (0.31); 2.32 (0.02) | | | | 2.31±8.28 (0.75); 3.08 (<0.01) | | | |
| ***Occupation** | | | | | | | | | | | | |
| ##Less Professional workers | 273 (67.4) | 45 (60.0) | 48 (92.3) | 7.98±7.84 | 312 (67) | 54 (83.1) | 0 | 4.78±3.65 | 237(69.9) | 77 (64.2) | 52 (71.2) | 9.66±8.49 |
| @Professional workers | 132 (32.6) | 30 (40.0) | 4 (7.7) | 5.97±5.05 | 154 (33) | 11 (16.9) | 1 (100) | 3.90±2.94 | 102(30.1) | 43 (35.8) | 21 (28.8) | 9.43±8.04 |
| X̅_d_ ± S.D. (S.E.); t (p) | 2.01± 7.09 (0.66); 3.05 (<0.01) | | | | 0.88± 3.44( (0.32); 2.75 (<0.01) | | | | 0.23± 8.35 (0.78); 0.29 (0.77) | | | |
| ***Education** | | | | | | | | | | | | |
| Less than Class 8 | 127 (31.4) | 26 (34.7) | 38 (73.1) | 9.79±8.96 | 152 (32.6) | 39 (60) | 0 | 5.48±3.92 | 107(31.6) | 46 (38.3) | 38 (52.1) | 11.24±9.11 |
| Class 8 or above | 278 (68.6) | 49 (65.3) | 14 (26.9) | 5.99±5.44 | 314 (67.4) | 26 (40) | 1 (100) | 3.96±3.05 | 232(68.4) | 74 (61.7) | 35 (47.9) | 8.67±7.74 |
| X̅_d_ ± S.D. (S.E.); t (p) | 3.8± 6.81 (0.62); 6.13 (<0.01) | | | | 1.52± 3.39 (0.31); 4.9 (<0.01) | | | | 2.57± 8.26 (0.75); 3.43 (<0.01) | | | |
| ***Income** | | | | | | | | | | | | |
| ≤ 36550 | 248 (61.2) | 38 (50.7) | 31 (59.6) | 7.07±7.04 | 281 (60.3) | 36 (55.4) | 0 | 4.38±3.26 | 212(62.5) | 67 (55.8) | 38 (52.1) | 9.14±7.83 |
| > 36550 | 157 (38.8) | 37 (49.3) | 21 (40.4) | 7.76±7.29 | 185 (39.7) | 30 (44.6) | 1 (100) | 4.71±3.74 | 127(37.5) | 53 (44.2) | 35 (47.9) | 10.26±9.03 |
| X̅_d_ ± S.D. (S.E.); t (p) | 0.69± 7.14 (0.63); 1.1 (0.27) | | | | 0.33± 3.46 (0.31); 1.06 (0.29) | | | | 1.12± 8.33 (0.74); 1.51 (0.13) | | | |
| ***Socio-economic status** | | | | | | | | | | | | |
| $ Middle to Upper class | 241 (59.5) | 44 (58.7) | 17 (32.7) | 6.47±6.21 | 275 (59) | 26 (40) | 1 (100) | 4.15±3.30 | 194(57.2) | 70  (58.3) | 38 (52.1) | 9.17±8.28 |
| ¶Lower class | 164 (40.5) | 31 (41.3) | 35 (67.3) | 8.50±8.08 | 191 (41) | 39 (60) | 0 | 4.98±3.63 | 145(42.8) | 50 (41.7) | 35 (47.9) | 10.14±8.41 |
| X̅_d_ ± S.D. (S.E.); t (p) | 2.03± 7.08 (0.62); 3.27(<0.01) | | | | 0.83± 3.44 (0.3); 2.77 (<0.01) | | | | 0.97± 8.34 (0.73); 1.33 (0.18) | | | |
| **Marital status** | | | | | | | | | | | | |
| Married | 384 (94.8) | 67 (89.3) | 37 (71.2) | 6.83±6.64 | 438 (94.0) | 49 (75.4) | 1 (100) | 4.30±3.30 | 320(94.4) | 104(86.7) | 64 (87.7) | 9.34±8.26 |
| ^Unmarried | 21 (5.2) | 8 (10.7) | 15 (28.8) | 13.16±9.59 | 28 (6.0) | 16 (24.6) | 0 | 6.82±4.38 | 19 (5.6) | 16 (13.3) | 9 (12.3) | 12.34±8.86 |
| X̅_d_ ± S.D. (S.E.); t (p) | 6.33± 6.93 (1.09); 5.81 (<0.01) | | | | 2.52± 3.4 (0.53); 4.75 (<0.01) | | | | 3± 8.31 (1.31); 2.29 (0.02) | | | |

#includes Janajati, Dalits, Madhesi, Muslim and Others (Giri);* classified according to Kuppuswamy scale; ##Unemployed, Unskilled worker, Semi-skilled worker, Skilled worker; @Arithmetic skill job, Semi-professional, Professional; ;$Upper, Upper Middle, Lower Middle; ¶Upper Lower,Lower^ includes Never married, Divorced, Widowed, Separated; ! X̅ d, mean difference; SD, standard deviation; SE, standard error of mean; t, t-statistic; p, p-value; p-value-significant at <.05.
